# Association of women’s empowerment with anaemia and haemoglobin concentration in children in sub-Saharan Africa: a multilevel analysis

**DOI:** 10.1101/2021.10.01.21264423

**Authors:** Calistus Wilunda, Milkah Wanjohi, Risa Takahashi, Elizabeth Kimani-Murage, Antonina Mutoro

## Abstract

**Background:** Childhood anaemia is an intractable public health problem, particularly in low- and middle-income countries. Women’s empowerment may improve child nutrition, but information on the relationship between different dimensions of women’s empowerment and childhood anaemia in sub-Saharan Africa (SSA) is limited. We assessed the associations between women’s empowerment and anaemia and haemoglobin (Hb) concentration among children in SSA.

**Methods:** We included 72 032 women and their singleton children aged 6-59 months from the most recent Demographic and Health Surveys conducted between 2006 and 2019 in 31 SSA countries. We constructed a four-dimensional women’s empowerment index using principal components analysis and used multilevel regression to assess the associations of these dimensions with child anaemia and Hb concentration.

**Results:** Of the included children, 65.8% were anaemic and the mean Hb concentration was 102.3 g/dl (SD 16.1). The odds of anaemia reduced with increasing empowerment in the dimensions of attitude towards violence [quintile (Q1) vs. Q5, OR 0.80; 95% CI 0.71–0.89, *P*_*tren*d_ <0.001], decision making (Q1 vs. Q5, OR 0.68; 95% CI 0.59–0.79, *P*_*trend*_ <0.001), education (Q1 vs. Q5, OR 0.80; 95% CI 0.72–0.89, *P*_*tre*nd_ <0.001), and social independence (Q1 vs. Q5, OR 0.89; 95% CI 0.79–1.00, *P*_*tren*d_ <0.015). Similarly, the mean Hb concentration increased with increasing women’s empowerment in the dimensions of attitude towards violence (Q1 vs. Q5, mean difference (MD) 0.96 g/dl; 95% CI 0.17–1.74, *P*_*tr*end_ = 0.009), decision making (Q1 vs. Q5, MD 0.73 g/dl; 95% CI 0.03–1.43, *P*_*trend*_ <0.001), social independence (Q1 vs. Q5, MD 1.65 g/dl; 95% CI 0.99–2.31, *P*_*trend*_ <0.001) and education (Q1 vs. Q5, MD 1.01 g/dl; 95% CI 0.50–1.52, *P*_*tre*nd_ <0.002).

**Conclusion:** Women empowerment was associated with reduced odds of anaemia and higher Hb concentration in children. Promotion of women empowerment may reduce the burden of childhood anaemia in SSA.

**Key questions:** *What is already known?:*

- Women empowerment may reduce maternal and child undernutrition, specifically wasting, stunting and Vitamin A deficiency.
- Women empowerment is a multidimensional concept and is often measured in an unstandardized way making it difficult to compare and interpret findings across studies.
- A survey-based women’s empowerment index (the SWPER index) was developed to measure women empowerment in a standardised manner across sub-Saharan African countries.

*What are the new findings?:*

- Children of women who were empowered in the attitude towards domestic violence, decision making, education, and social independence domains had higher haemoglobin concentrations and were less likely to be anaemic than those of less empowered women.
- Empowerment in decision making seemed to have the strongest association with anaemia.

*What do the new findings imply?:*

- Promotion of women empowerment in attitude towards violence, decision making, social independence and education domains may contribute towards reducing the burden of anaemia among sub-Saharan African children.

## Introduction

Childhood anaemia is a major intractable public health problem affecting 42% of the children aged between 6 and 59 months world-wide.^1^ Sub-Saharan Africa (SSA) has one of the highest prevalence in the world. In 2016, 60% of children in SSA were anaemic.^1^ Anaemia is an established risk factor for mortality in low- and middle-income countries and its detrimental consequences on development and human potential have long been recognised.^2 3^ To effectively prevent anaemia, a better understanding of its social determinants is required.

The underlying causes of anaemia include diseases such as hookworm infestation, sickle cell disorders, thalassemias, schistosomiasis and malaria^4 5^ and inadequate nutrient intake, which leads to micronutrient deficiencies such as iron deficiency. These underlying causes can be attributed to poor child care and nutrition practices, which are linked to lack of maternal education, large family sizes and lack of autonomy, all of which are aspects of women empowerment – defined as the process of increasing women’s power and autonomy so that they have control over strategic life choices that allows them to fully realize their potential.^6 7 8^

Women empowerment may reduce maternal and child undernutrition, specifically wasting, stunting and Vitamin A deficiency,^9-12^ but a lack of a standardized measure and definition of this multidimensional concept makes comparison and interpretation of study findings difficult.^9^ Moreover, whereas several studies have assessed the association between women empowerment and wasting or stunting in children, only a few have comprehensively assessed the association between women empowerment and child anaemia in SSA.

To mitigate the problem of measuring women empowerment and to allow for comparisons across studies, a survey-based women’s empowerment index (the SWPER index) was developed and validated using data from the Demographic and Health Survey (DHS) from 34 African countries.^13^ This opened opportunities to explore the association of women empowerment with a range of health and nutritional indicators in women and children using a standardized measure. We aimed to assess the association between women’s empowerment – based on the SWPER index – and anaemia among children aged between 6 and 59 months in SSA.

## Methods

### Data source and study population

This study utilised pooled data from the most recent Demographic and Health Surveys (DHS) conducted between 2006 and 2019 in 31 countries in SSA. All countries with data on haemoglobin measurement were included (supplementary Table 1). The detailed methodology of DHS is available elsewhere.^14^ In brief, DHS are cross-sectional nationally-representative household surveys conducted in low- and middle-income countries to provide data for a wide range of population, health, and nutrition indicators. Standard DHS have a sample sizes usually between 5,000 and 30,000 households and are typically conducted about every 5 years. Households are selected through stratified multistage cluster sampling method and socio-demographic, health and nutrition data are collected mainly from women aged 15-49 years, and children below five years using interviewer administered questionnaires. Blood samples are collected from women and children in all households or in a random subset of selected households based on considerations such as the required sample size and financial costs.

The study population for the current analysis is women and their singleton children aged 6-59 months. We restricted the analysis to the youngest child and to singletons to avoid clustering of children at the household level. The initial pooled dataset contained 323,505 children and their mothers. From this number, we excluded the following: children from households not selected for haemoglobin (Hb) measurement (n = 138,210), non-usual residents (n = 3,792), missing Hb data (n = 48,608), not youngest child (n = 43,083), twins (n = 1589), unmarried women (n = 13,036), missing data to compute the empowerment index (n = 3,155).

### Variables

The outcome variables were standardized measures of child Hb concentration, adjusted for altitude, and anaemia. ^15^ Anaemia was defined as Hb concentration <11.0 grams/decilitre (g/dl).^16^ Hb measurement involved collection of blood samples from children aged 6-59 months using a microcuvette from a drop of blood taken from a finger or heel prick. Hb analysis was conducted on site using a portable HemoCue® photometer. This method of Hb measurement is efficient and compares well with the standard laboratory methods.^17^

The exposure was SWPER index for women’s empowerment. We constructed a SWPER index on the pooled dataset as previously described by Ewerling *et al*.^13^ This involved identifying the indicators of women empowerment available across the surveys and recoding them as shown in supplemental Table 2. Next, we performed principal component analysis of the selected indicators and generated a scree plot to determine the number of components to retain. Using the conventional Eigen value of >1, we retained four components that loaded well on variables related to attitude towards domestic violence (beating justified if wife neglects children, refuses to have sex with husband, burns food), decision making (decision on health care and purchase of household goods), social independence (age at cohabitation and age at first birth) and education (reading newspapers and the difference in the years of schooling between the wife and husband) (Supplemental Figure 1 and Supplemental Table 3). Women were then ranked into empowerment quintiles based on the factor scores of each of the retained components, with the lowest quintile representing the least empowered and the highest quintile representing the most empowered.

The following were considered as covariates: year of the survey, parity, wealth index quintile, place of residence, living with partner, woman’s age (continuous), child’s age (continuous) and child’s sex. We re-constructed the wealth index on the combined dataset using principal component analysis of several proxy wealth index indicators including type of housing material, asset ownership and access to utilities.^18^

### Statistical analysis

Our analysis was based on a pooled dataset of all the included countries. We used descriptive statistics to summarise participants’ characteristics and women’s empowerment variables. We then ran three-level mixed effects logistic and linear regression models to assess the associations of women empowerment with child anaemia and haemoglobin concentration. We quantified the associations using odds ratios (OR, for anaemia) and mean differences (MD, for Hb) with 95% confidence intervals (CIs). In the multilevel analyses, households were nested within clusters and clusters were nested within countries. The analysis accounted for the survey design and sample weights. As multilevel analysis of survey data requires specification of weights at different levels, we generated country level weights by dividing the population of the respective county at the time of the survey by the sample size of each country.^19^ For the cluster-level weights, we used the survey weight variable (which is a product of the household-level weight and the cluster-level weight) supplied in the DHS datasets. We performed both unadjusted and adjusted analyses. In adjusted analyses, all dimensions of women empowerment were mutually adjusted for each other and for the covariates above. However, for social independence, we did not adjust for woman’s age and parity as these are potential mediators. We assessed for linear trends by including the empowerment variable as a continuous variable in the models. There was no evidence of multi-collinearity based on variance inflation factors (VIFs): VIFs ranged from 1.01 to 1.93, with a mean of 1.64. All analyses were performed using Stata 15 and *P* values <0.05 were considered statistically significant.

### Ethical considerations

Country-specific DHS protocols were approved by relevant ethics committees in each country and respondents provided informed consent. DHS data were accessed with permission from the DHS Program. Ethical review was not required for this study because de-identified open-access datasets were used in the analysis.

## Results

This study included 72 032 (unweighted) mother-child dyads. Of the included children, 48 653 (65.8%, weighted) were anaemic and the mean Hb concentration was 102.3 [standard deviation (SD) 16.1]. The mean age of the mothers was 30.0 (SD 7.1) years and 34.2% had a parity of 1-2 with 24.8% having a parity of >6 (Table 1). Most of the women were from rural areas (67.7%) and married (85.2%). The mean age of children was 25.1 (SD 13.9) and 48.9% of them were female.

**Table 1.** Distribution of study participants from 31 SSA countries by selected socio-demographic characteristics

| Characteristic | Urban (N=21 360) | Rural (N = 50 672) | Total (N = 72 032) |
| --- | --- | --- | --- |
| Woman's age, mean (SD) | 30.2 (6.5) | 29.9 (7.3) | 30.0 (7.1) |
| Parity |  |  |  |
| 1-2 | 43.4 | 30.4 | 34.2 |
| 3-4 | 18.3 | 15.5 | 16.3 |
| 5-6 | 23.2 | 25.2 | 24.6 |
| >6 | 15.1 | 28.9 | 24.8 |
| Wealth index quintile |  |  |  |
| Poorest | 2.1 | 27.6 | 20.1 |
| Poorer | 3.8 | 27.0 | 20.2 |
| Middle | 9.0 | 24.6 | 20.0 |
| Richer | 29.3 | 16.0 | 20.0 |
| Richest | 55.7 | 4.7 | 19.7 |
| Missing data | 0.1 | 0.1 | 0.1 |
| Year of interview |  |  |  |
| 2006-2008 | 8.3 | 24.0 | 19.4 |
| 2009-2011 | 5.6 | 7.7 | 7.1 |
| 2012-2014 | 17.7 | 17.3 | 17.4 |
| 2015-2017 | 30.4 | 27.4 | 28.3 |
| 2018-2019 | 38.0 | 23.6 | 27.8 |
| Marital status |  |  |  |
| Married | 81.2 | 86.9 | 85.2 |
| Living with partner | 18.8 | 13.2 | 14.8 |
| Sex of household head |  |  |  |
| Male | 85.1 | 88.3 | 87.3 |
| Female | 14.9 | 11.7 | 12.7 |
| Child's sex |  |  |  |
| Male | 51.8 | 50.8 | 51.1 |
| Female | 48.2 | 49.2 | 48.9 |
| Child's age, mean (SD) | 26.0 (14.4) | 22.7 (13.7) | 25.1 (13.9) |

Table 2 summarises the distribution of study subjects according to indicators of women empowerment. Among the women, the following reasons justified wife beating: going out without telling husband (35.1%), neglecting children (37.7%), arguing with husband (35.8%), refusing to have sex with husband (30.8%), and burning food (22.7%). One third (33.4%) of women were from households where the husband/partner or other person was the sole decision maker on their healthcare while 43.5% reported that their husband/partner was the usual decision maker on large household purchases (Table 2). Most of the women (84.4%) never read newspapers and 61.2% were currently working. The women’s age at first co-habitation was 18.2 (SD 4.2) years and the mean age at first birth was 19.3 (SD 3.8) years. Compared to their partners, on average, the women were 8.0 (SD 7.1) years younger and had schooled for 1.3 (SD 3.8) years less. Overall, the proportions of women who felt wife beating was justified for any of the mentioned reasons or whose husband/partner or other person was the main decision maker were significantly higher among the anaemic than the non-anaemic children. Woman’s age at first cohabitation, age at first birth and the age difference with husband were significantly higher among the anaemic than non-anaemic children. The proportion of women who read a newspaper least once a week or were currently working was significantly lower among the anaemic than the non-anaemic children (Table 2).

**Table 2:** Percent distribution of women in 31 SSA countries according to indicators of women empowerment

| Empowerment indicator | Total<sup>1</sup><br>(N = 72 032) | Child anaemic<sup>1</sup> |  | P value |
| --- | --- | --- | --- | --- |
|  |  | No (N = 23 379) | Yes (N = 48 653) |  |
| Beating justified if: |  |  |  |  |
| Wife goes out without telling husband |  |  |  | <0.001 |
| Yes | 35.1 | 32.7 | 36.3 |  |
| Don't know | 0.6 | 0.4 | 0.6 |  |
| No | 64.4 | 66.9 | 63.1 |  |
| Wife neglects children |  |  |  | <0.001 |
| Yes | 37.6 | 35.6 | 38.7 |  |
| Don't know | 0.5 | 0.5 | 0.5 |  |
| No | 61.9 | 64.0 | 60.8 |  |
| Wife argues with husband |  |  |  | <0.001 |
| Yes | 35.8 | 32.9 | 37.3 |  |
| Don't know | 0.5 | 0.5 | 0.5 |  |
| No | 63.7 | 66.7 | 62.2 |  |
| Wife refuses to have sex with husband |  |  |  | <0.001 |
| Yes | 30.8 | 28.2 | 32.2 |  |
| Don't know | 0.7 | 0.9 | 0.7 |  |
| No | 68.5 | 71.0 | 67.2 |  |
| Wife burns food |  |  |  | 0.002 |
| Yes | 22.7 | 21.1 | 23.5 |  |
| Don't know | 0.4 | 0.3 | 0.4 |  |
| No | 77.0 | 78.6 | 76.1 |  |
| Who usually decides on respondent's health care |  |  |  | <0.001 |
| Husband/partner or other alone | 38.9 | 30.9 | 43.0 |  |
| Respondent and husband/partner | 45.5 | 51.2 | 42.5 |  |
| Respondent alone | 15.7 | 17.9 | 14.5 |  |
| Who usually decides on large household purchases |  |  |  | <0.001 |
| Husband/partner or other alone | 43.5 | 35.8 | 47.6 |  |
| Respondent and husband/partner | 46.1 | 53.1 | 42.4 |  |
| Respondent alone | 10.4 | 11.1 | 10.0 |  |
| Who usually decides on visits to family/relatives |  |  |  | <0.001 |
| Husband/partner or other alone | 33.4 | 27.1 | 36.6 |  |
| Respondent and husband/partner | 50.4 | 56.9 | 47.0 |  |
| Respondent alone | 16.3 | 16.1 | 16.4 |  |
| Age at first cohabitation | 18.2 (4.2) | 18.5 (4.2) | 18.0 (4.2) | <0.001 |
| Age at first birth | 19.3 (3.8) | 19.5 (3.8) | 19.2 (3.8) | <0.001 |
| Woman's minus husband's age | -8.0 (7.1) | -7.5 (6.5) | -8.3 (7.5) | <0.001 |
| Years of schooling |  |  |  | <0.001 |
| None | 40.6 | 33.2 | 44.5 |  |
| 1-4 | 12.9 | 12.8 | 13.0 |  |
| 5-8 | 25.2 | 27.6 | 23.9 |  |
| ≥8 | 21.3 | 26.5 | 18.6 |  |
| Woman's minus husband's years of schooling | -1.3 (3.8) | -1.3 (3.7) | -1.3 (3.8) | 0.875 |
| Frequency of reading newspaper |  |  |  | <0.001 |
| Not at all | 84.4 | 80.6 | 86.3 |  |
| Less than once a week | 10.0 | 12.3 | 8.8 |  |
| At least once a week | 5.6 | 7.1 | 4.9 |  |
| Worked in the past 12 months |  |  |  | <0.001 |
| No | 31.5 | 29.0 | 32.8 |  |
| In the past year | 7.3 | 8.1 | 6.8 |  |
| Currently working | 61.2 | 62.9 | 60.4 |  |

<sup>1</sup>Values are expressed as percent or mean (SD)

The prevalence of anaemia was highest among the least empowered and lowest among the most empowered women across all the four dimensions of empowerment (Table 3). The highest prevalence (76.4%) was observed among those least empowered in the decision-making dimension and the lowest prevalence was among those most empowered educationally (58.4%). Unadjusted regression results showed that the odds of anaemia reduced linearly with increasing level of empowerment. Similarly, the mean Hb concentration generally increased with increasing level of empowerment across all the four dimensions of empowerment (Table 3).

**Table 3:** Unadjusted associations of dimensions of women empowerment with anaemia and haemoglobin concentration in children from 31 SSA countries

| Empowerment index quintile (Q) | Anaemia |  |  | Haemoglobin concentration, g/dl |  |  |
| --- | --- | --- | --- | --- | --- | --- |
|  | Percent | OR (95% CI) | P-value | Mean (SD) | MD (95% CI) | P-value |
| Attitude towards violence |  |  |  |  |  |  |
| Q1 | 69.7 | 1 (ref.) |  | 100.6 (15.1) | 1 (ref.) |  |
| Q2 | 66.6 | 0.87 (0.80-0.94) | 0.001 | 101.9 (16.3) | 1.00 (0.48-1.52) | <0.001 |
| Q3 | 67.7 | 0.89 (0.80-1.00) | 0.059 | 101.4 (17.5) | 1.05 (0.36-1.75) | 0.003 |
| Q4 | 64.0 | 0.77 (0.66-0.89) | 0.001 | 103.0 (16.5) | 2.20 (1.35-3.05) | <0.001 |
| Q5 | 60.9 | 0.69 (0.57-0.84) | 0.001 | 104.4 (15.0) | 3.61 (2.08-5.13) | <0.001 |
| P<sub>trend</sub> |  | 0.001 |  |  | <0.001 |  |
| Decision making |  |  |  |  |  |  |
| Q1 | 76.4 | 1 (ref.) |  | 98.2 (17.1) | 1 (ref.) |  |
| Q2 | 70.0 | 0.74 (0.68-0.81) | <0.001 | 100.7 (16.2) | 1.14 (0.34-1.93) | 0.005 |
| Q3 | 62.1 | 0.56 (0.49-0.63) | <0.001 | 103.5 (14.8) | 2.84 (2.11-3.58) | <0.001 |
| Q4 | 59.9 | 0.51 (0.44-0.58) | <0.001 | 104.7 (14.9) | 3.73 (2.91-4.54) | <0.001 |
| Q5 | 60.4 | 0.51 (0.43-0.59) | <0.001 | 104.3 (16.0) | 3.13 (2.01-4.25) | <0.001 |
| P<sub>trend</sub> |  | <0.001 |  |  | <0.001 |  |
| Social independence |  |  |  |  |  |  |
| Q1 | 70.0 | 1 (ref.) |  | 99.9 (16.1) | 1 (ref.) |  |
| Q2 | 67.0 | 0.90 (0.81-0.99) | 0.037 | 101.6 (16.1) | 1.42 (0.51-2.33) | 0.002 |
| Q3 | 66.6 | 0.90 (0.76-1.05) | 0.168 | 102.3 (16.2) | 1.67 (1.08-2.25) | <0.001 |
| Q4 | 63.6 | 0.79 (0.68-0.92) | 0.003 | 103.1 (16.5) | 2.43 (1.40-3.47) | <0.001 |
| Q5 | 61.6 | 0.73 (0.59-0.90) | 0.005 | 104.4 (15) | 3.92 (2.27-5.57) | <0.001 |
| P<sub>trend</sub> |  | 0.015 |  |  | <0.001 |  |
| Educational empowerment |  |  |  |  |  |  |
| Q1 | 67.8 | 1 (ref.) |  | 101.5 (15.7) | 1 (ref.) |  |
| Q2 | 71.4 | 1.17 (0.95-1.44) | 0.133 | 99.5 (18.0) | -1.12 (-2.43-0.19) | 0.094 |
| Q3 | 69.4 | 1.07 (0.99-1.17) | 0.090 | 100.6 (16.4) | -1.04 (-2.02- -0.05) | 0.039 |
| Q4 | 61.7 | 0.80 (0.71-0.91) | 0.001 | 104.4 (14.9) | 1.98 (1.31-2.65) | <0.001 |
| Q5 | 58.4 | 0.70 (0.62-0.80) | <0.001 | 105.4 (14.4) | 2.97 (1.96-3.98) | <0.001 |
| P<sub>trend</sub> |  | <0.001 |  |  | <0.001 |  |

Women in Q1 are the least empowered while those in Q5 are the most empowered.
OR: Odds ratio; MD: Mean difference

The results of multivariable analysis (Table 4) show that the odds of anaemia reduced with increasing empowerment in the dimensions of attitude towards violence (Q1 vs. Q5, OR 0.80; 95% CI 0.71–0.89, *P*_*tren*d_ <0.001), decision making (Q1 vs. Q5, OR 0.68; 95% CI 0.59–0.79, *P*_*trend*_ <0.001), education (Q1 vs. Q5, OR 0.80; 95% CI 0.72–0.89, *P*_*tre*nd_ <0.001), and social independence (Q1 vs. Q5, OR 0.89; 95% CI 0.79–1.00, *P*_*tren*d_ <0.015). Similarly, the mean Hb concentration increased with increasing women’s empowerment in the dimensions of attitude towards violence (Q1 vs. Q5, mean difference (MD) 0.96 g/dl; 95% CI 0.17–1.74, *P*_*tr*end_ = 0.009), decision making (Q1 vs. Q5, MD 0.73 g/dl; 95% CI 0.03–1.43, *P*_*trend*_ <0.001), social independence (Q1 vs. Q5, MD 1.65 g/dl; 95% CI 0.99–2.31, *P*_*trend*_ <0.001) and education (Q1 vs. Q5, MD 1.01 g/dl; 95% CI 0.50–1.52, *P*_*tre*nd_ <0.002).

**Table 4:** Adjusted associations of dimensions of women empowerment with anaemia and haemoglobin concentration in children from 31 SSA countries

| Empowerment index quintile (Q) | Anaemia |  | Haemoglobin concentration, g/dl |  |
| --- | --- | --- | --- | --- |
|  | OR (95% CI) | P-value | MD (95% CI) | P-value |
| Attitude towards violence |  |  |  |  |
| Q1 | 1 (ref.) |  | 1 (ref.) |  |
| Q2 | 0.90 (0.79-1.02) | 0.103 | 0.36 (-0.26-0.98) | 0.261 |
| Q3 | 0.93 (0.85-1.02) | 0.121 | 0.11 (-0.64-0.85) | 0.778 |
| Q4 | 0.85 (0.77-0.93) | 0.001 | 0.79 (0.21-1.37) | 0.007 |
| Q5 | 0.80 (0.71-0.89) | <0.001 | 0.96 (0.17-1.74) | 0.017 |
| P<sub>trend</sub> | <0.001 |  | 0.009 |  |
| Decision making |  |  |  |  |
| Q1 | 1 (ref.) |  | 1 (ref.) |  |
| Q2 | 0.82 (0.76-0.88) | <0.001 | 0.33 (-0.21-0.88) | 0.231 |
| Q3 | 0.69 (0.61-0.78) | <0.001 | 1.35 (0.80-1.89) | <0.001 |
| Q4 | 0.63 (0.53-0.74) | <0.001 | 1.76 (0.94-2.57) | <0.001 |
| Q5 | 0.68 (0.59-0.79) | <0.001 | 0.73 (0.03-1.43) | 0.042 |
| P<sub>trend</sub> | <0.001 |  | <0.001 |  |
| Educational empowerment |  |  |  |  |
| Q1 | 1 (ref.) |  | 1 (ref.) |  |
| Q2 | 1.13 (0.96-1.34) | 0.134 | -0.79 (-1.69-0.12) | 0.088 |
| Q3 | 1.03 (0.94-1.13) | 0.550 | -0.73 (-1.68-0.21) | 0.128 |
| Q4 | 0.84 (0.77-0.91) | <0.001 | 0.93 (0.21-1.64) | 0.011 |
| Q5 | 0.80 (0.72-0.89) | <0.001 | 1.01 (0.50-1.52) | <0.001 |
| P<sub>trend</sub> | <0.001 |  | 0.002 |  |
| Random effects variance |  |  |  |  |
| Country, estimate (SE) | 0.00 (0.00) |  | 19.95 (21.12) |  |
| Cluster, estimate (SE) | 0.52 (0.06) |  | 0.00 (0.00) |  |
| Social independence |  |  |  |  |
| Q1 | 1 (ref.) |  | 1 (ref.) |  |
| Q2 | 0.92 (0.78-1.07) | 0.265 | 1.15 (0.18-2.12) | 0.020 |
| Q3 | 0.95 (0.86-1.06) | 0.335 | 1.08 (0.56-1.61) | <0.001 |
| Q4 | 0.88 (0.80-0.97) | 0.016 | 1.30 (0.74-1.86) | <0.001 |
| Q5 | 0.89 (0.79-1.00) | 0.047 | 1.65 (0.99-2.31) | <0.001 |
| P<sub>trend</sub> | 0.015 |  | <0.001 |  |
| Random effects variance |  |  |  |  |
| Country, estimate (SE) | 0.00 (0.00) |  | 18.74 (23.41) |  |
| Cluster, estimate (SE) | 0.52 (0.07) |  | 0.00 (0.00) |  |

Women in Q1 are the least empowered while those in Q5 are the most empowered.
OR: Odds ratio; MD: Mean difference
<sup>1</sup>All variables, except social dependence, were mutually adjusted for each other and for survey year, parity, wealth index, place of residence, living with partner, interview year, woman's age, child's age and child's sex. Social dependence was adjusted for all the variables except woman's age and parity as these may be mediators.

## Discussion

This study examined the association between women empowerment and childhood anaemia in SSA. We found that women who were empowered in the attitude towards domestic violence, decision making, education, and social independence domains were less likely to have anaemic children.

Our study findings are consistent with the existing literature on the role of women empowerment in child nutrition in SSA.^20-23^ However, studies on women empowerment and child nutrition in this region have focused mainly on anthropometrical indicators, particularly stunting and underweight, ^20^ with only a few looking at anaemia.^23 24^ One study, whose aim was to explore pathways by which women’s empowerment influences child nutritional status,^24^ was based on DHS data from five East African countries and found significant and direct positive associations between women’s instrumental agency in household decisions and child haemoglobin concentrations.^24^ However, the study did not find a direct link between intrinsic agency (attitudes toward violence) with anemia.^24^ The study also found a significant positive association, both direct and mediated through instrumental agency, between assets (social empowerment) and anemia.^24^ Our findings are consistent with those of a study in India, which found that mother’s ability to make decisions about own health care, contribution to the family income, newspaper reading or negative attitude towards domestic violence increased the odds that her child will not have anaemia.^25^ Associations between poor iron status and women’s empowerment in agriculture have also been reported.^26^

Women’s empowerment can prevent anaemia in children through poverty reduction, improved access to food and promotion of awareness on nutrition, and promotion of appropriate child feeding and other care practices. Moreover, empowerment can lead to better health seeking behaviours ^10^ including treatment of infections such as malaria, iron supplementation, and deworming, which are key interventions to reduce anaemia.

The prevalence of childhood anaemia was highest among women who were least empowered especially in the decision-making dimension. In a study in Ethiopia, higher women’s empowerment in household decision making was associated with reduced anaemia in children.^23^ Women participation in decision-making regarding household purchases can promote child nutrition and reduce anaemia though improved dietary diversity.^27^ Financial empowerment is also likely to lead to improved child and household nutrition because compared to men, women are more likely to invest a larger proportion of their income in their families.^28^ Education and decision making go hand-in-hand because educated women are likely to access gainful employment and contribute to household finances and decision making. They are also more likely to have knowledge on appropriate child care practices.

This is the first time the association between the SWPER index and anaemia and Hb concentration in children in SSA has been assessed. The findings from this study therefore add to the growing evidence of the importance of women empowerment in promoting nutrition in children. The study’s large sample size increased the power to detect significant differences while the inclusion of data from a large number of countries from SSA increases the generalizability of our findings. Despite these strengths, this being a cross-sectional study, we cannot infer causation or establish a temporal relationship between the indicators of empowerment and the study outcomes. Moreover, our assessment of women empowerment was limited by the variables measured across all the included countries.

In conclusion, women empowerment was associated with reduced odds of anaemia and higher Hb concentration in children. Promotion of women empowerment in attitude towards violence, decision making, social independence and education dimensions may contribute towards reducing the burden of anaemia among children in SSA.

## Data Availability

Data are available in a public, open access repository. Data for this study were sourced from Demographic and Health surveys (DHS) and available here: http://dhsprogram.com/data/available-datasets.cfm.

## Contributors

CW and RT contributed to the study design and conceptualisation. CW performed the analysis. CW, MW and AM drafted the initial draft. EK-M, RT critically reviewed the manuscript for its intellectual content. All authors read and amended drafts of the paper and approved the final version. CW had final responsibility to submit for publication.

## Patient consent for publication

No consent to publish was needed for this study as we did not use any details, images or videos related to individual participants. In addition, data used are available in the public domain.

## Ethics approval and consent to participate

The DHS survey protocols and procedures are approved by the ethics committee of ORC Macro and partner organisations of participating countries.

