## Supplementary file for "Association of women’s empowerment with anaemia and haemoglobin concentration in children in sub-Saharan Africa: a multilevel analysis"

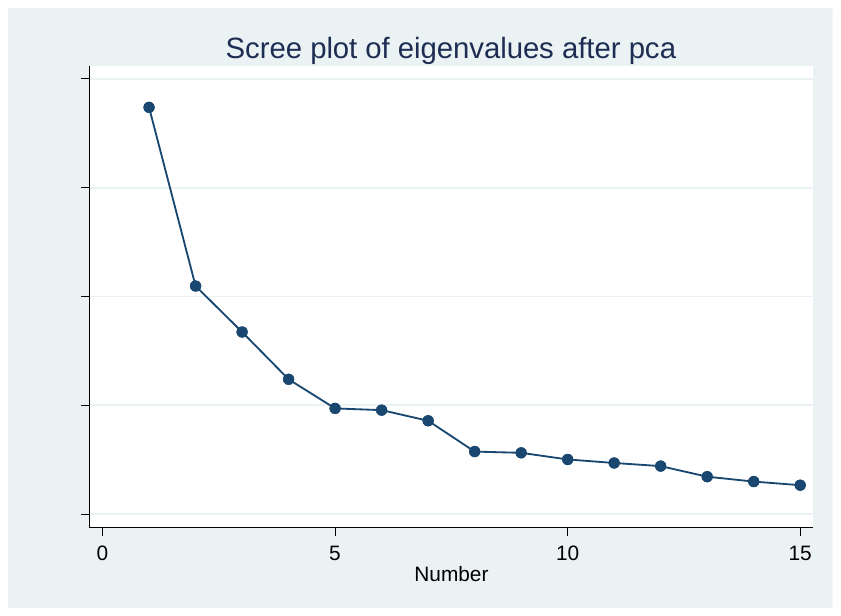

**Supplemental Figure 1: Scree plot showing the four extracted components with Eigenvalues >1**

**Supplemental Table 1: Included countries**

|  | Country | Survey year |
| --- | --- | --- |
| 1 | Angola | 2015/16 |
| 2 | Burkina Faso | 2010 |
| 3 | Benin | 2011/12 |
| 4 | Burundi | 2016/17 |
| 5 | DRC | 2013/14 |
| 6 | Code d'Ivoire | 2011 |
| 7 | Cameroon | 2018 |
| 8 | Ethiopia | 2008 |
| 9 | Gabon | 2012 |
| 10 | Ghana | 2014 |
| 11 | Gambia | 2013 |
| 12 | Guinea | 2018 |
| 13 | Lesotho | 2014 |
| 14 | Madagascar | 2008/09 |
| 15 | Mali | 2018 |
| 16 | Malawi | 2015/16 |
| 17 | Mozambique | 2011 |
| 18 | Nigeria | 2018 |
| 19 | Niger | 2012 |
| 20 | Namibia | 2013 |
| 21 | Rwanda | 2014 |
| 22 | Sierra Leone | 2013 |
| 23 | Senegal | 2017 |
| 24 | Sao Tome | 2008/09 |
| 25 | Swaziland | 2006/07 |
| 26 | Togo | 2013/14 |
| 27 | Tanzania | 2015/16 |
| 28 | Uganda | 2016 |
| 29 | South Africa | 2016 |
| 30 | Zambia | 2018 |
| 31 | Zimbabwe | 2015 |

**Supplemental Table 2: Coding of the items used in the development of the women’s empowerment index**

| Variable | Code or unit |
| --- | --- |
| Beating not justified if wife goes out without telling husband | Justified=–1; don’t know=0; not justified =1 |
| Beating not justified if wife neglects children | Justified=–1; don’t know=0; not justified =1 |
| Beating not justified if wife argues with husband | Justified=–1; don’t know=0; not justified =1 |
| Beating not justified if wife refuses to have sex with husband | Justified=–1; don’t know=0; not justified =1 |
| Beating not justified if wife burns food | Justified=–1; don’t know=0; not justified =1 |
| Who usually decides on respondent's health care | Husband or other alone=–1; joint=0;  respondent alone=1 |
| Who usually decides on large household purchases | Husband or other alone=–1; joint=0;  respondent alone=1 |
| Who usually decides on visits to family or relatives | Husband or other alone=–1; joint=0;  respondent alone=1 |
| Years of schooling | Years |
| Worked in the past 12 months | No=0; in the past year=1;  have a job, but on leave past 7 days=2;  currently working=2 |
| Frequency of reading newspaper | Not at all=0; <once a week=1;  ≥once a week=2 |
| Age at first cohabitation | Years |
| Age at first birth | Years |
| Difference in years of schooling: woman’s minus husband’s years of schooling | Years |
| Age difference: woman’s minus husband’s age | Years |

Source: Ewerling *et al^13^*

**Supplemental Table 3: Principal component analysis factor loadings based on the combined dataset including all study countries**

| Variable | Attitude towards violence | Decision making | Social independence | Educational empowerment |
| --- | --- | --- | --- | --- |
| Beating not justified if wife goes out without telling husband | **0.4563** | -0.0063 | -0.0047 | 0.0072 |
| Beating not justified if wife neglects children | **0.4664** | -0.0216 | -0.0156 | -0.0135 |
| Beating not justified if wife argues with husband | **0.4606** | 0.0119 | -0.0010 | -0.0015 |
| Beating not justified if wife refuses to have sex with husband | **0.4377** | 0.0185 | 0.0012 | 0.0142 |
| Beating not justified if wife burns food | **0.4071** | -0.0143 | 0.0093 | -0.0045 |
| Person who usually decides on respondent's health care | -0.0033 | **0.5645** | -0.0278 | 0.0401 |
| Person who usually decides on large household purchases | -0.0116 | **0.5644** | -0.0274 | 0.0139 |
| Person who usually decides on visits to family or relatives | 0.0100 | **0.5415** | -0.0249 | -0.0258 |
| Years of schooling | 0.0390 | 0.0899 | 0.1027 | **0.5895** |
| Worked in the past 12 months | -0.0187 | 0.1537 | **0.1669** | -0.1868 |
| Frequency of reading newspaper | 0.0006 | 0.0319 | 0.0230 | **0.5595** |
| Age at first cohabitation | 0.005 | 0.0004 | **0.6674** | 0.034 |
| Age at first birth | -0.0093 | -0.046 | **0.6818** | -0.0099 |
| Difference in years of schooling: woman’s minus husband’s years of schooling | -0.0498 | -0.1117 | -0.1088 | **0.548** |
| Age difference: woman’s minus husband’s age | 0.0109 | 0.1456 | **0.1907** | 0.0111 |

Note: This table was obtained after varimax rotation
